# A Foundational Exome Resource for Jordan: Regional Ancestry, Novel Variants, and Implications for Clinical Variant Interpretation

**DOI:** 10.64898/2026.05.23.26353895

**Authors:** Tawfiq Froukh

## Abstract

**Background:** The genetic architecture of Middle Eastern populations is underrepresented in global genomic databases, which increases the rate of Variants of Uncertain Significance (VUSs) and complicates clinical variant interpretation.

**Methods:** Whole exome sequencing was conducted on 90 healthy Jordanians and analysed 66 genetically unrelated individuals. Principal Component Analysis (PCA), ADMIXTURE and f4 statistics were used to infer ancestry against 1000 Genomes and Human Genome Diversity Project reference populations, including Palestinian, Bedouin, Druze, and Mozabite. The identified variants were annotated against seven population databases and against ClinVar and were tested for enrichment with one sided Fisher’s exact tests and Benjamini-Hochberg false discovery rate (FDR) correction.

**Results:** A total of 201,577 variants were identified at a median target depth of 71X. PCA analysis clustered the Jordanian individuals with the Palestinian, Bedouin, and Druze reference populations. The Jordanian profile exhibited a sub-Saharan African ancestry component (median of 7.2%), which is comparable to Bedouin and Palestinian references, and four individuals carried more than 30%. The f4 statistics did not show African affinity excess in the remaining individuals relative to Palestinians. A total of 3,281 variants (1.6%) had no records in any of the seven population databases or in ClinVar. They include 153 variants with high-impact effect, ten of them lay in constrained genes. The cohort had 108 ClinVar pathogenic or likely pathogenic variants, including eight absent from population databases, seven of them lay in genes associated with recessive diseases.

**Conclusion:** The study provides first regional resource of exome variation in Jordan. The value sits in documenting ancestry and variants that may help interpretation when combined with larger, ancestry-matched datasets.

## INTRODUCTION

Historically, the Middle East region was a path for human migration among Africa, Europe and Asia [1]. Despite the central role of the Middle East in understanding the patterns of human migration and its high prevalence of genetic diseases, it remains underrepresented in global genomic databases [2]. For example, recent analysis of the Genome-Wide Association studies (GWAS) reveals only 0.96% of participants of Arab descent [3]. Furthermore, the Middle Eastern genomes make up only 0.2% of the total samples in the Genome Aggregation Database (gnomAD) [4]. This shortage of Middle Eastern data in public databases limits the generalizability of genomic research and triggers global health inequities [5].

The genetic landscape in Jordan is complex because it is characterized by significant admixture events [6, 7]. For example, Levantine populations harbour varying degrees of African ancestry (4%-15%) with an average mixture date of 72 generations ago [8]. These unique signals build and accumulate genetic variations which are considered as population-specific variants (PSVs). Even the current regional databases did not reveal all of the PSVs because they focus on the Arabian Peninsula or the Middle East at large. Consequently, identifying these PSVs is essential to uncover the “long-tail” of rare variants that formulate the genetic architecture in Jordan [9]. The present-day genome studies of Middle-Eastern populations, including Jordanians, describe a demographic history with several admixture and drift events [10], and regional resources such as Qatari [11] and Turkish [12] genomes have begun to fill the gap.

Severe clinical consequences would probably result from the underrepresentation of Jordan genomic variants [13]. In European-centric reference databases, rare PSVs are misclassified as pathogenic or as variant of uncertain significance (VUS) [14, 15]. Recent genomic studies show that population-specific databases can facilitate the reclassification of high-impact variants previously thought to be pathogenic [16]. Therefore, targeted sequencing of underrepresented populations is needed to refine clinical annotations and minimize or even prevent diagnostic errors [17, 18].

Using the whole-exome sequencing (WES) data from a healthy Jordanian cohort, this study aims to (i) describe the ancestry of the cohort relative to regional reference populations, (ii) catalogue the variants and quantify those absent from public databases, (iii) test under conservative conditions whether the variants are enriched relative to public databases, and (iv) report ClinVar-listed pathogenic variants carried by healthy individuals and discuss the limits of using small cohort for clinical variant interpretation.

## MATERIALS AND METHODS

Ethical approval for this study was obtained from the Institutional Review Board (IRB) of Philadelphia University in Jordan IRB number (2/1/2024-2025). Healthy adult volunteers (n=90; aged 18-25 years) from the Department of Biotechnology and Genetic Engineering at Philadelphia University in Jordan were recruited according to the criteria in Table 1. All participants provided written informed consent to publish the results of their exomes anonymously.

**Table 1.** Recruitment criteria for participants.

| **Individual information** | **Criteria** |
|---|---|
| Age | Between 18 and 25 years |
| Ethnicity | Jordanian |
| Sex | 1:1 ratio |
| Blood pressure | Normal |
| Disease history and physical disability: cardiovascular, nephrological, urological, nervous system, gastrointestinal, endocrine and metabolic, blood, connective tissue, and cancer | Unremarkable |

Peripheral blood samples were collected from all participants by trained medical technicians at Philadelphia University in Jordan. Total genomic DNA was extracted using the QIAGEN FlexiGene DNA kit (QIAGEN, Hilden, Germany) according to the manufacturer’s protocol.

Exomes were captured using Agilent SureSelect Human All Exon Kits (V5, V6, V7, and V8). The sequences were obtained as paired-end reads on the Illumina NovaSeq 6000 at two sequencing facilities. The raw data output was approximately 9 Gb per sample. Target sizes differed between designs (35.1 to 60.5 Mb). Mosdepth [19] was used to calculate mean depth and the percentage of target bases cover at 10X, 20X and 30X over the target regions of the design for each individual.

All bioinformatics analysis was conducted using the pipeline implemented in Philadelphia University. The generated paired-end reads were trimmed using Trimmomatic-0.39 and aligned using BWA to GRCh38.p14 [20, 21]. Picard was used to mark duplicates and ABRA was used to realign indels [22, 23]. Variants were called for each individual with BCFtools mpileup (minimum mapping and base quality 20, and maximum depth 2,000), and BCFtools call in gVCF mode, and were left-aligned and normalized [24]. Functional variants were annotated using SnpEff against the GRCh38 MANE (Matched Annotation from NCBI and EMBL-EBI) version 1.5 [25, 26].

Only high-quality genotypes were maintained for statistical analysis including a raw read depth (DP ≥ 10), phred-scaled quality score (QUAL ≥ 30), average mapping quality (MQ ≥ 40), phred-scaled genotype quality (GQ ≥ 20), phred-scaled strand bias P value (SP ≤ 60), read position bias (−5 ≤ RPBZ ≤ 5), mapping quality versus strand bias (−5 ≤ MQSBZ ≤ 5), variant allele fraction (VAF ≥ 0.2) and soft clipping fraction (SCR/DP ≤ 0.3). To minimize technical artefacts, stringent filtering was conducted by excluding variants overlapping with UCSC Segmental Duplications, RepeatMasker regions and the ENCODE Blacklist. The maintained variants were grouped into four subsets: high, moderate, low and modifier putative impacts based on the sorting system adopted by SnpEff for multiple annotation effect [25]. Variants were annotated using SnpSift [26] with the following public resources: OMIM (genemap2 generated 5/9/2026) [27, 28], ClinVar (release of 22/8/2026) [29], dbSNP (build 157)[30], 1000 Genomes Project [31], TOPMed freeze 8 [32], gnomAD joint (exome and genome) v4.1, including its Middle Eastern genetic ancestry group (gnomAD_mid) and gene constrained metrics (loss-of-function observed/expected upper bound fraction, LOEUF; probability of loss-of-function intolerance, pLI) [4, 33], ALFA total (release 4, 20241028) [34], RegeneronME (around 983,578 exomes) [35], and the Qatari Genome [11]. Variants statistics were acquired using BCFtools.

Variants were assigned to six groups according to their presence in the above resources: A, no record in any public resource (novel); B, a rsID; C, a ClinVar entry but no population record; D, population records present but all allele counts zero; E, allele count above zero in some but not all population resources (TOPMed, ALFA, gnomAD including gnomAD_mid, 1000 Genomes, RegeneronME, and Qatari); and F, allele count above zero in all population resources.

King-robust [36] in PLINK2 [37–40] was used to estimate relatedness using autosomal common variants. Maximal set of unrelated individuals were used for all population-level analysis by removing related individuals based on a kinship coefficient above 0.0442. The cohort was merged with 1000 Genome Phase 3 individuals [31] (Africans, Europeans and South Asians) and with Human Genome Diversity Project (HGDP) individuals including Bedouins, Palestinian, Druze, and Mozabite [41]. Only informative variants were maintained based on: autosomal chromosomes, variants having minor allele frequencies (MAF) ≥ 0.05, missing rate (--geno) ≤ 0.02 and the exclusion of variants showing extreme deviation from Hardy-Weinberg Equilibrium (HWE; *p* < 10^-6^). Linkage disequilibrium (LD) pruning was conducted using –indep-pairwise with a 50-variant window, a 5-variant step size and a threshold of 0.2 in order to ensure PC capturing broad ancestral rather than localized LD. The LD-pruned dataset (11,972 variants) was used to calculate PCA [39] using the command –pca which generated the top 10 eigenvectors and eigenvalues. ADMIXTURE [42] was used to estimate Model-based ancestry for k = 2 to 8 with five-fold cross-validation on the 11,972 pruned variants in 1,124 individuals: all Jordanians and HGDP and 50 individuals from each of the 1000 Genomes populations. A sensitivity PCA was conducted using all 1000 Genomes populations, including the American and East Asian populations, together with HGDP and the cohort (3,027 individuals; 12,289 LD-pruned variants using the same filters; S1 Fig). For f4 statistics, ADMIXTOOLS 2 [43] was used on all 20,915 variants without pruning because a 5-Mb jackknife blocks account for linkage between nearby variants, in 268 individuals from seven populations (Jordanian, Palestinian, Bedouins, Druze, Mozabite, Mandenka, and French). All PCA plots were conducted with R and association of each principal component with capture design was tested within Kruskal-Wallis test [44].

In the unrelated cohort, allele counts (AC) and allele numbers (AN) were compared with those of each population resource. Variants without frequency records in all population resources, including all novel variants were only described and not tested. Other variants were tested if they were in at least 80% of alleles (AN ≥ 106), had an allele count in all populations resources, and had a highest frequency across population resources below 1%. The targeted comparator (Index) was the resource with the highest allele frequency, chosen as the reference with least favorable to enrichment; comparison was also repeated without Qatari genome, whose allele counts were reconstructed from the rounded frequencies (minimum 0.46%), and against the gnomAD_mid and the Qatari Genome individually. For the hypothesis concerned an excess of frequency, one-tailed Fisher’s exact test was used to identify variants that are significantly enriched in the studied dataset (over-represented) relative to the global databases, together with a zero-truncated binomial test that conditions on the variant have been observed [45]. To control for expected high rate of false positives associated with testing thousands of variants, *p-*values were adjusted for multiple comparisons using Benjamini-Hochberg False Discovery Rate (FDR) procedure [46]. Statistical significance is defined as an FDR-adjusted *p*-value < 0.05; the number of tests is reported for each comparator. The analyzed variants are these with at least two alleles (primary) or one allele (sensitivity), and the analysis was repeated after excluding individuals with an African component above 30%. Allele frequency confidence intervals are exact (Clopper-Pearson) [47]. All analyses are implemented in Python (v3.10.18) utilizing the scipy.stats library for the exact tests and the statsmodels.stats.multitest module for FDR correction [48].

## RESULTS

Of the 90 sequenced individuals, 66 form the analysis cohort based on the kinship coefficient < 0.0442. Across the 66 individuals, the median depth over the target was 71X, and a median of 91.7% of target bases was covered at 20X or more (Table 2). The depth varies between designs, from a mean of 56X in the V6 at one facility to 109X in V7. Two individuals had low coverage (13X and 22X, with 17% and 48% of the target bases at 20X or more); their uncertain genotypes were set to missing by the filters. The per-individual median transition/transversion ratio was 2.89 and the median ratio of heterozygous to non-reference homozygous genotypes per individual was 1.69.

**Table 2.** Capture designs, sequencing facilities and coverage of the 66 unrelated individuals.

| **Capture kit and facility** | **n** | **Target (Mb)** | **Mean depth (X)** | **Mean % target ≥20X** |
|---|---|---|---|---|
| SureSelect V5, facility GeJo | 5 | 58.8 | 83.8 | 92.2 |
| SureSelect V6, facility GeJo | 17 | 50.1 | 74.3 | 85.8 |
| SureSelect V6, facility Jo | 25 | 60.5 | 56.1 | 86.9 |
| SureSelect V7, facility GeJo | 18 | 49.3 | 109.1 | 96.8 |
| SureSelect V8, facility Jo | 1 | 35.1 | 66.1 | 97.5 |
| All individuals (median; range) | 66 | - | 71.1 (13.0-137.3) | 91.7 (17.0-98.1) |

### Ancestry in a regional context

PCA revealed a cluster of the Jordanian individuals with the Palestinian, Druze, and Bedouin populations laying between the European and African clusters with a few Jordanian individuals extending toward the African and Mozabite populations along PC1 (Fig 1A and 1B; PC1 68.2%, PC2 11.9%, PC3 5.9% of variance).

**Figure 1.**
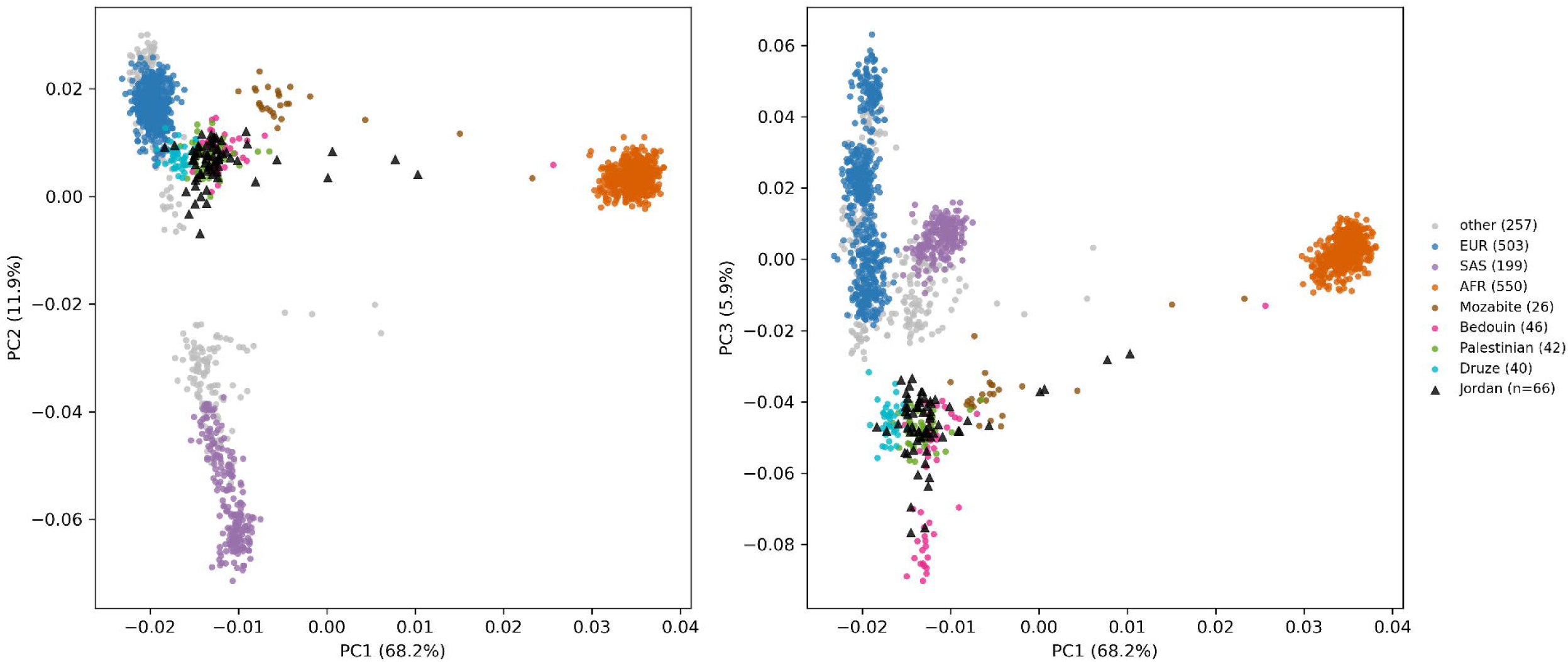
Ancestry of the Jordanian cohort in a regional context. (A) PC1 versus PC2 and (B) PC1 versus PC3 of 1,729 individuals: the 66 unrelated Jordanian individuals (black triangles), 1000 Genomes African (AFR), European (EUR) and South Asian (SAS) populations, and HGDP populations including Bedouin, Palestinian, Druze and Mozabite. (C) ADMIXTURE at K = 4; each bar is one individual, the Jordanian individuals are sorted by their African component and colours are labelled by the reference population in which each component is highest. PC3 was associated with capture design and is shown.

The cross-validation error in ADMIXTURE, was lowest (0.5034) at K = 4 and almost identical for K = 4 to K = 6. The dominated component in the Jordanian profile at K = 4 peaks in the Bedouin reference, with smaller components in the European and South Asian references and a Sub-Saharan African component with a median of 7.2% (90^th^ percentile 15.8 %, maximum 51.7%), similar to Bedouin (7.7%) and Palestinian (6.2%) references (Fig 1C). At every K from 2 to 8, four Jordanian individuals had an African component above 30%.

In a sensitivity PCA, adding the American and East Asian populations, there was no indication of affinity between the Jordanian individuals and the American populations (S1 Fig). The Jordanian individuals remained in the Middle Eastern reference (Palestinian, Bedouin, and Druze), while the American population extended along a different axis, toward the East and South Asian clusters on PC2 and below the European on PC3.

In the f4 analysis, the Jordanian individuals did not show excess African affinity relative to the Palestinian individuals (f4 = −4.1 x 10^-5^, SE 1.1 x 10^-4^, Z = −0.36) except four Jordanians, which were with the highest African component (f4 = 0.0067, SE 2.9 x 10^-4^, Z =23.2).

For all 66 Jordanian individuals, PC3 was associated with capture design (Kruskal-Wallis P = 0.004), and (Kruskal-Wallis P = 0.045) after excluding the four individuals with the highest African component and the two low-coverage individuals, whereas PC1 and PC2 were not (Kruskal-Wallis P > 0.25).

### Overview of the Exome dataset

The total number of high-quality variants identified in the 66 individuals is 201,577. The dataset is dominated by Single Nucleotide Polymorphisms (SNPs, n=187,663) with minor insertions and deletions (Indels, n=13,914). The observed transition/transversion ratio across all 187,663 SNPs is 2.69, lower than the per-individual median because of rare variants, and both values are consistent with the expected range for exome data. Approximately 39% of the dataset are singletons and 141,394 variants were called in at least 80% of alleles. Single variant density is not reported because the capture designs vary in their target size from 35.1 to 60.5 Mb.

### Distribution of functional impact

According to SnpEff and MANE transcripts, the majority of the variants had a low predicted impact (n=55,558), followed by moderate (n=50,141), and variants classified with high impact accounts for 0.77% of the total dataset (n=1,559). A further 94,319 variants are classified as modifiers, residing in non-coding regions without predicted primary effect on protein structure (Table 3, Fig 2A). The proportion of variants without any public record (group A) was highest among high-impact variants (153 of 1,559; 9.8%) compared to moderate (1,343 of 50,141; 2.7%), low (665 of 55,558; 1.2%) and modifier variants (1,120 of 94,319; 1.2%).

**Figure 2.**
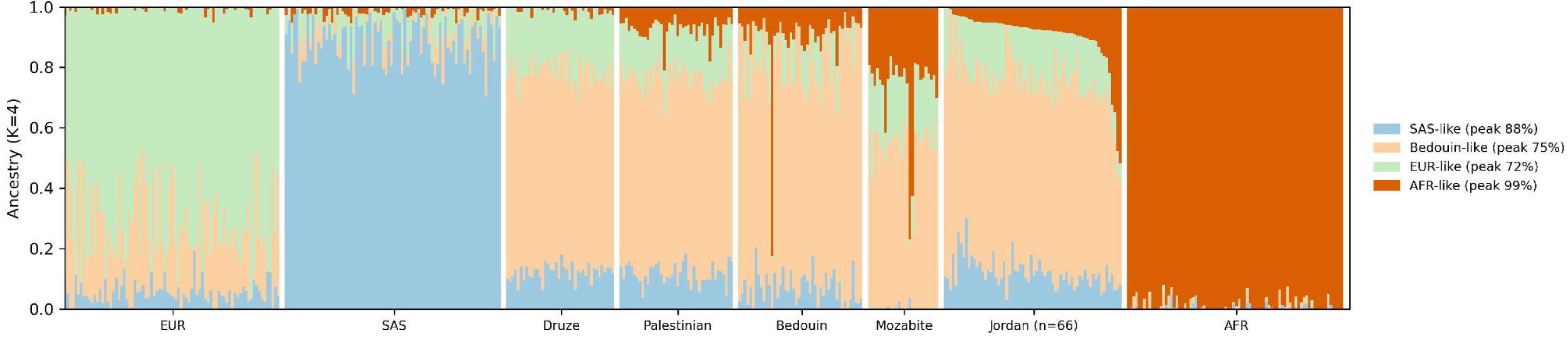
Variants in the 66 unrelated individuals. (A) Number of variants by predicted impact (SnpEff, MANE 1.5), with and without a public record (group A, no record in any population database or ClinVar and no rsID); log scale. (B) Variants by presence in public reference databases (groups A to F, defined in the Methods); log scale.

**Table 3.** Variants in the 66 unrelated individuals by predicted impact and ppresence in public databases.

| **Impact** | **A novel** | **B rsID only** | **C ClinVar only** | **D counts zero** | **E some** | **F all seven** | **Total** |
|---|---|---|---|---|---|---|---|
| High | 153 | 2 | 9 | 10 | 1,122 | 263 | 1,559 |
| Moderate | 1,343 | 117 | 97 | 191 | 24,781 | 23,612 | 50,141 |
| Low | 665 | 51 | 32 | 136 | 23,609 | 31,065 | 55,558 |
| Modifier | 1,120 | 92 | 5 | 352 | 40,152 | 52,598 | 94,319 |
| **Total** | **3,281** | **262** | **143** | **689** | **89,664** | **107,538** | **201,577** |

### Variants without population records

A total number of 3,281 variants (1.6%) had no records in any population database or ClinVar and no rsID (group A, novel), 262 had an rsID only, 143 had a ClinVar entry but no population record, and 689 had records with all alleles counts equal to zero. The remaining variants had a count above zero in some (89,664; 44.5%) or all seven (107,538; 53.4%) population databases (Table 3, Fig 2B).

### Enrichment relative to public databases

None of the 1,798 testable variants (at least two alleles) and of the 7,285 testable variants (with at least one allele) had significant enrichment after FDR correction against the Index comparator. However, many variants appeared enriched against single regional databases: 4,330 of 13,949 tests against gnomAD_mid and 711 of 7,241 against the Qatari Genome (FDR < 0.05, at least two alleles). By excluding the Qatari Genome from the index, 22 variants (3 to 7 alleles) had FDR < 0.05, 19 of them were against gnomAD_mid, but none had FDR < 0.05 when variants with a single allele were included (7,285 tests). The same pattern was obtained by excluding the four individuals carrying more than 30% African ancestry: 0 of the 1,488 tests against the Index; 21 against the Index without Qatari, and none when single alleles were included.

None of the 19 variants reported as enriched in the preprint (doi: 10.64898/2026.05.23.26353895) remained significant against the Index comparator (smallest FDR 0.40; 0.10 with at least two alleles). Four of them were significant against gnomAD_mid (FDR < 0.05; range 0.03-0.05) after excluding the Qatari Genome from the index and only variants with at least two alleles were tested; none of the four was significant with the zero-truncated test. Moreover, the preprint analyses were based on 90 individuals and a reclassification was proposed for the variants in ECE2 and THPO (allele frequency 3.89%; 7 of 180 alleles) and in IL1RN (allele frequency 3.93%; 7 of 178 alleles). After removing related individuals (kinship coefficient > 0.0442; n=66) the frequencies fell to 3.79% for ECE2 and THPO (5 of 132 alleles; exact 95% CI 1.2-8.6%) and to 2.3% in IL1RN (3 of 130 alleles). The ECE2 variant (c.2236C>T;p.Arg746Trp; VUS in ClinVar) and the THPO variant (c.183G>T;p.Leu61Leu; Conflicting in ClinVar) were carried by the same five individuals, consistent with the same haplotype. Their frequency was 8.2-fold higher that Qatari Genome (0.46%; OR 9.2; one-sided P = 5.0 x 10^-4^) and 11.5-to 12.1-fold higher than in gnomAD_mid (0.33% and 0.31%; OR 12.7 and 13.4; P = 1.5 x 10^-4^ and 1.3 x 10^-4^). While both remained significant against each of these two databases after FDR correction (FDR 0.012 and 0.007, respectively), none of them remained significant against the Index comparator (FDR 0.1). The IL1RN variant was not significant against the Index comparator (FDR 0.31). For the Qatari, counts were reconstructed and are not an independent observation. Therefore, the proposed reclassifications in the preprint are not supported by the present analysis.

### Clinical annotation and pathogenicity potential

Of the 153 high-impact novel variants (group A), 151 were singletons and 124 were frameshift or stop-gain (93 frameshift and 31 stop-gained). Of these 153 variants, 133 were retained using a stricter support criterion other than the genotype filters (carrier depth ≥ 20 and variant allele fraction between 0.3 and 0.7). Of these retained variants, ten lay in genes with LOEUF < 0.6; from which only one gene (DST) has a recessive OMIM phenotype (Table 4). All these ten variants are singletons in a single heterozygous individual and have not been validated by an independent method.

**Table 4.**
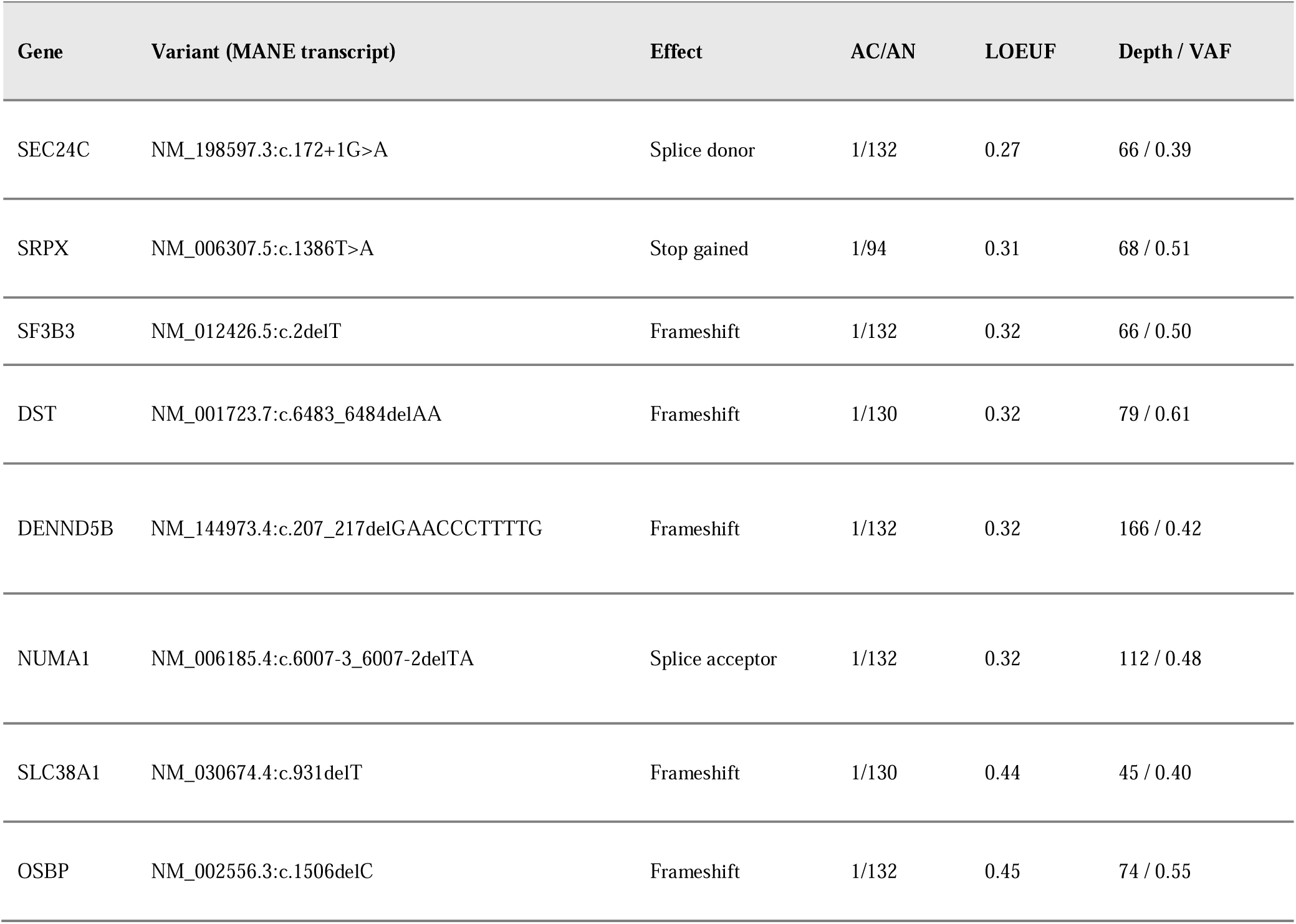
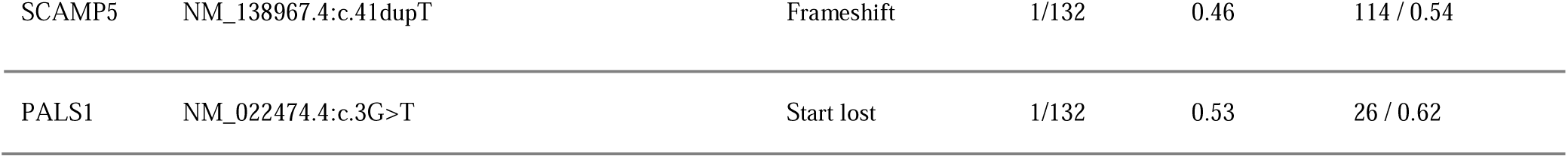
Candidate loss-of-function variants without population or ClinVar records in genes with LOEUF < 0.6 (carrier depth ≥ 20 and variant allele fraction 0.3-0.7).

| Gene | Variant (MANE transcript) | Effect | AC/AN | LOEUF | Depth / VAF |
|---|---|---|---|---|---|
| SEC24C | NM_198597.3:c.172+1G>A | Splice donor | 1/132 | 0.27 | 66 / 0.39 |
| SRPX | NM_006307.5:c.1386T>A | Stop gained | 1/94 | 0.31 | 68 / 0.51 |
| SF3B3 | NM_012426.5:c.2delT | Frameshift | 1/132 | 0.32 | 66 / 0.50 |
| DST | NM_001723.7:c.6483_6484delAA | Frameshift | 1/130 | 0.32 | 79 / 0.61 |
| DENND5B | NM_144973.4:c.207_217delGAACCCTTTTG | Frameshift | 1/132 | 0.32 | 166 / 0.42 |
| NUMA1 | NM_006185.4:c.6007-3_6007-2delTA | Splice acceptor | 1/132 | 0.32 | 112 / 0.48 |
| SLC38A1 | NM_030674.4:c.931delT | Frameshift | 1/130 | 0.44 | 45 / 0.40 |
| OSBP | NM_002556.3:c.1506delC | Frameshift | 1/132 | 0.45 | 74 / 0.55 |
| SCAMP5 | NM_138967.4:c.41dupT | Frameshift | 1/132 | 0.46 | 114 / 0.54 |
| PALS1 | NM_022474.4:c.3G>T | Start lost | 1/132 | 0.53 | 26 / 0.62 |

In the 66 individuals, 108 variants are listed in ClinVar as pathogenic or likely pathogenic and 143 variants with no population records but with ClinVar entry. Of the 143 variants, 35 were classified as benign, 1 as benign/likely benign, 4 as conflicting, 95 as VUS, and 8 as pathogenic or likely pathogenic (Table 5). Each of the 8 variants was observed once. In which, seven variants lie in genes associated with autosomal recessive disorders in OMIM (CDC45, CFAP300, COL25A1, KATNB1, MSH3, VWA3B and WRN), and one in gene associated with both dominant and recessive deafness (CEACAM16).

**Table 5.** ClinVar pathogenic or likely pathogenic variants without population records.

| Gene | Variant (MANE transcript) | ClinVar classification | AC/AN | OMIM phenotype (first listed) |
|---|---|---|---|---|
| CDC45 | NM_003504.5:c.165G>A | Pathogenic | 1/132 | Meier-Gorlin syndrome 7 |
| CEACAM16 | NM_001039213.4:c.662-1G>C | Pathogenic | 1/130 | Deafness, autosomal dominant 4B |
| CFAP300 | NM_032930.3:c.693_694insGA | Pathogenic | 1/132 | Ciliary dyskinesia, primary, 38 |
| COL25A1 | NM_198721.4:c.1198G>A | Likely pathogenic | 1/130 | Fibrosis of extraocular muscles, congenital, 5 |
| KATNB1 | NM_005886.3:c.1A>G | Pathogenic | 1/132 | Lissencephaly 6, with microcephaly |
| MSH3 | NM_002439.5:c.3046G>T | Pathogenic | 1/132 | Familial adenomatous polyposis 4 |
| VWA3B | NM_144992.5:c.1191T>G | Pathogenic/Likely pathogenic | 1/132 | Spinocerebellar ataxia, autosomal recessive 22 (provisional) |
| WRN | NM_000553.6:c.2952_2953delAT | Pathogenic/Likely pathogenic | 1/132 | Werner syndrome |

## DISCUSSION

This is a descriptive study for the exome variation in 66 unrelated Jordanian adults, in which their ancestry is placed among the regional reference populations. Furthermore, the study quantifies the novel variants compared to seven population resources and to ClinVar, and tests the enriched variants relative to the population resources. Three main findings were obtained. First, resemblance to Levantine and Arabian reference populations, with four individuals carry African ancestry. Second, 1.6% of the total variants are novel which are mostly high-impact predicted variants. Third, no significant enriched variant is identified against the most conservative comparator, although the significant enrichment of thousands against single regional databases.

The ancestry analysis in this study is consistent with the regional structure described before for the eight Middle Eastern populations [10]. The ADMIXTURE results demonstrate a dominant Bedouin-like component with smaller European and South Asian components. This aligns with their finding that Bedouins lie close to ancient Levantines and that present-day Levantines are drawn toward Bronze Age Europeans [10]. In addition, the same study reported more African ancestry in Arabians than in Levantines, which agrees with this study: Bedouin reference (7.7%) is slightly above Palestinian reference (6.2%) and the Jordanian median (7.2%) lies between them, although this difference is small and was not tested. The estimate of African admixture within the last 2,000 years, mostly 500 to 1,000 years ago [10] is compatible with the modest and fairly uniform African fraction with no excess relative to Palestinians (f4 Z = −0.36). It cannot account for the four individuals with more than 30% African ancestry (f4 Z = 23.2). Because 66 exomes cannot support admixture dating, no claim about timing in this study is established. Also, the four individuals cannot be related to the sub-communities described in earlier Jordanian studies [6, 7], because participants reported Jordanian ethnicity only (Table 1).

PC3 association can be interpreted in the scope of technical component from the capture kits. This is because PC3 was associated with capture design (P = 0.004, and P = 0.045 after excluding the four individuals with high African ancestry and the two low-coverage samples), whereas PC1 and PC2 were not (P > 0.25).

The negative results of enrichment tests against the Index comparator are interpreted by the conservative design of the comparison and by the statistical power. With 66 individuals, it is unlikely for a variant with two or three alleles to survive FDR correction across thousands of tests. The significant enrichment against gnomAD_mid or Qatari genome alone is not evidence of Jordan-specific enrichment, but the 22 variants obtained after removing the Qatari Genome are kept for future follow-up in larger cohort.

The variants proposed for reclassification in the preprint (doi: /10.64898/2026.05.23.26353895) illustrate the need for unrelated samples and conservative comparators. The apparent enrichment from the 90 individuals was not significant against the Index comparator in the 66 unrelated individuals. According to the American College of Medical Genetics and Genomics/Association for Molecular Pathology (ACMG/AMP) guidelines [49], standalone benign classification (BA1) requires allele frequency above 5%, and BS1 requires a frequency greater than expected for the disorder, in a large ancestry-matched cohort [50, 51]; 132 alleles cannot provide this.

The novel variants in the cohort (1.6% based on exome for all 66 individuals) are less than regional programs such as the Qatari resource [52], which records 1.7% novel variants per individual genome. Part of this difference is due to exome (in this study) versus genome (in Qatari). Nevertheless, these variants show that Jordanian individuals carry variants unseen in global databases. The work published under the Middle East Variation was built to capture genetic variation in the Middle East to rescue the underrepresentation in most comprehensive genomic databases and to improve annotation [9]. The novel variants in this study add a Levantine cohort to MEV’s picture, since Jordan has low representation compared to the Arabian Peninsula. The higher fraction of high-impact novel variants (9.8%) compared to others (2.7% of moderate-impact and 1.2% of low-impact and modifier variants) is expected because selection keeps high-impact variants at low frequency, which means higher probability to be unrecorded in public databases. However, calling artefacts possibly accumulated in high-impact class is also possible but cannot be separated from the selection effect without orthogonal validation.

The ten loss-of-function candidates in constrained genes, including the one in *DST* (Table 4) are candidates that require confirmation by Sanger sequencing. The identified loss-of-function variants, and the identified pathogenic or likely pathogenic ClinVar variants are expected for recessive conditions in healthy carriers [53]. Because these variants are in genes associated with recessive disorders, their presence does not contradict their classification. Late-onset disease or reduced penetrance cannot be excluded based on a healthy status at 18-25 years. Moreover, no clinical phenotyping beyond the observed criteria was performed and therefore, no conclusion about misclassification can be drawn. The heterozygous pathogenic variant in the gene *CEACAM16* is recorded in ClinVar with autosomal recessive hearing loss whereas heterozygous carriers have normal hearing [54].

The study has the following limitations: small sample size with 132 alleles the lowest frequency is 0.76%, four different capture designs in two sequencing facilities, allele counts derived from the rounded frequencies in Qatari, and the power of exome data for demographic inference.

## CONCLUSION

The exome data from 66 unrelated Jordanian place them within the Levantine and Arabian reference groups and documents variants not previously reported in public databases. The data cannot establish allele frequencies precisely neither support clinical reclassification. However, the data may help to improve variant interpretation when integrated with larger ancestry-matched datasets.

## Data Availability Statement

Findings summary are deposited in Zenodo (DOI: 10.5281/zenodo.24464548). It includes allele counts and annotations for all variants in the 66 individuals, enrichment analyses, novelty groups, candidate variant tables, coverage and quality metrics per-individual identified only by anonymous codes, PCA coordinates and ADMIXTURE proportions of the cohort, and the software versions and parameters.

## Acknowledgments

The author extends his thanks to all anonymous participants and to the lab technician Mrs. Eman Al-bsoul for her assistance extracting DNA samples.

## Author Contributions

**TF:** Conceptualization, Methodology, Software, Validation, Formal analysis, Investigation, Resources, Data curation, writing – original draft, Writing – review & editing, Visualization, Project administration.

## Supporting information

**S1 Fig.**
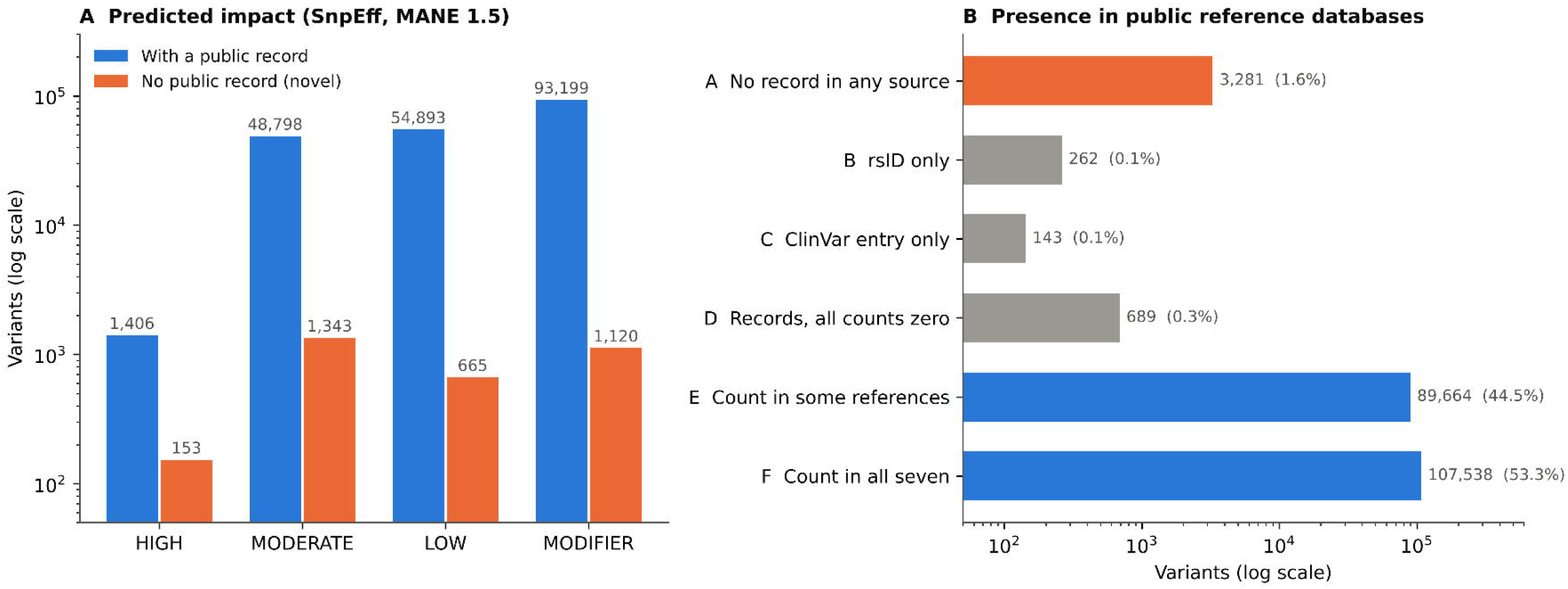
Sensitivity PCA with all 1000 Genomes populations. PC1 versus PC2 (left) and PC1 versus PC3 (right) of 3,027 individuals: the 66 unrelated Jordanian individulas (black triangles), all 1000 Genomes populations grouped as African (AFR), American (AMR), East Asian (EAS), European (EUR) and South Asian (SAS), and HGDP populations including Bedouin, Palestinian, Druze and Mozabite. The same LD-pruned variants as in Fig. 1 were used.

